# Machine learning-based equations for improved body composition estimation in Indian adults

**DOI:** 10.1101/2024.10.17.24315678

**Authors:** Nick Birk, Bharati Kulkarni, Santhi Bhogadi, Aastha Aggarwal, Gagandeep Kaur Walia, Vipin Gupta, Usha Rani, Hemant Mahajan, Sanjay Kinra, Poppy AC Mallinson

## Abstract

Bioelectrical impedance analysis (BIA) is commonly used as a lower-cost measurement of body composition as compared to dual-energy X-ray absorptiometry (DXA) and magnetic resonance imaging (MRI) in large-scale epidemiological studies. However, existing equations for body composition based on BIA measures may not generalize well to all settings.

We combined BIA measurements (TANITA BC-418) with skinfold thickness, body circumferences, and grip strength to develop equations to predict six DXA-measured body composition parameters in a cohort of Indian adults using machine learning techniques. The participants were split into training (80%, 1297 males and 1133 females) and testing (20%, 318 males and 289 females) data to develop and validate the performance of equations for total body fat mass (kg), total body lean mass (kg), total body fat percentage (%), trunk fat percentage (%), L1-L4 fat percentage (%), and total appendicular lean mass (kg), separately for males and females. Our novel equations outperformed existing equations for each of these body composition parameters. For example, the mean absolute error for total body fat mass was 1.808 kg for males and 2.054 kg for females using the TANITA’s built-in estimation algorithm, 2.105 kg for males and 2.995 kg for females using Durnin-Womersley equations, and 0.935 kg for males and 0.976 kg for females using our novel equations.

These equations may provide improved estimation of body composition in research and clinical contexts conducted in India and will be made available as an online application for use in future research in these populations.

## Background

Measurements of body composition, such as body fat percentage, can provide meaningful health-related insights. Total body fat and intra-abdominal fat have been previously associated with metabolic syndrome^1^, while lean body mass can be protective against chronic diseases and age-related adverse conditions^2,3^. Further, there is some evidence that these relationships are particularly pronounced in adults of South Asian ethnicities^1,2^, underscoring the importance of accurately recording these body composition metrics in epidemiological studies. While many studies of individuals of South Asian ethnicities use metrics such as overall weight and body mass index (BMI), these metrics may fail to distinguish between fat mass and lean mass, and therefore provide an incomplete assessment of body composition^4,5^. Thus, there is clinical and public health relevance in more accurately, and separately, characterizing both fat and lean components of body weight in South Asian populations. It should be noted that the label of ‘South Asian’ is used throughout this manuscript to refer to people who identify as having ancestral origin from countries in the South Asian region (defined by the World Bank as India, Pakistan, Bangladesh, Sri Lanka, Nepal, Bhutan and the Maldives) and may share tradition, diet, and values. By using this terminology, we do not mean to imply that any such differences are genetic in origin, or that South Asians, or Indians, represent a single ethnic or cultural group.

Bioelectrical impedance analysis (BIA) is commonly used as a feasible, lower-cost measurement of body composition in population-based studies as compared to dual-energy X-ray absorptiometry (DXA) and magnetic resonance imaging (MRI)^6^. Commercially and clinically available BIA devices, such as TANITA body composition analyzers, utilize inbuilt equations to estimate total and segmental lean and fat mass from a combination of body segment impedance values, sex, height, and weight^7^. However, these prediction equations were generally developed and validated in predominately European ethnic populations and may not generalize to other ethnic groups^8,9^. A study of 200 healthy Asian Indian individuals found that in-built equations based on white ethnic populations from a TANITA analyzer underestimated total body fat percentage by 8.3%, or by 5.4% when using equations fit based on individuals of Japanese ethnicity, compared with DXA-derived values^10^. A study of 39 Indian adults found that BIA methods underestimated fat mass as compared to four-compartment model, suggesting population-specific equations are needed^11^. Another study of healthy Indian adult men found that existing BIA equations tended to underestimate body fat compared with deuterium oxide dilution as a reference method^12^, while studies among Indian children have reported both under– and over-estimation of body fat percentage as compared to DXA-derived values^13–15^. Validation studies have previously demonstrated that impedance analysis led to underestimation of total body water, a component of lean mass, in Indian adults^16^.

The relative inaccuracy of BIA-based body composition estimation in Asian Indians compared with white populations may be related to the observed differences in body composition between White and Asian Indian adults^17^. In particular, Indian men and women have been found to have higher body fat percentage than their European ethnic counterparts of the same BMI^18–20^. The findings of Rush et al suggest that this difference may be related to the presence of higher abdominal fat mass and lower appendicular muscle mass among Asian Indian adults as compared to European, Maori, and Pacific Island ethnic adults after adjustment for age, height, and weight^18^. Further, a study of multiple community-based cohorts in the United States found that adults of South Asian ethnicity tended to have less lean mass overall than adults of other ethnic backgrounds^21^. Consequently, there is a relatively high prevalence of “normal weight obesity” in India^22,23^. The precise mechanisms responsible for these differences remain subject to debate, though may relate to exposure to common environmental factors^24^. Nonetheless, because of these differences in the presentation of adiposity, there is a need for better methods to estimate body composition in Asian Indian populations^17^. We are aware of one study that developed a novel BIA-based equation to predict DXA-derived body fat among young adult Indian males^25^, and another that developed lean body mass equations in people of Indian Asian ethnicity living in New Zealand^17^, but both had relatively small samples. Furthermore, no Indian-specific equations are available for estimation of central body fat, which may be particularly relevant for understanding cardiometabolic risks in Indian adults.

While BIA-based methods are increasingly popular for measuring body composition in research and clinical contexts, other prediction equations for body composition based on simple physical measurements have been previously developed. For example, the Durnin-Womersley equations are frequently used in epidemiological studies for estimating body fat percentage from skinfold measurements^26^. However, studies also suggest these equations perform less well in Asian (and other non-white) ethnic groups compared with white populations^27^ and indicate the potential of such anthropometric measures to further improve body composition estimation equations through recalibration to specific populations^28^.

The aim of the current work is to develop novel algorithms to predict DXA-derived fat and lean mass metrics calibrated for an adult Indian population. We use the BIA-based default predictions from the TANITA BC-418 (a widely used segmental body composition analyser) as a performance baseline, and then incorporate additional predictors from the TANITA output alongside potentially relevant anthropometric measures related to body stature, adiposity, and muscularity (namely age, standing height, circumferences, skinfolds, and grip strength). We evaluate the performance of the model against DXA-derived body composition metrics in a held-out sample. We explore various combinations of predictors and prediction methods, including penalized regression and non-parametric machine learning algorithms. The best-performing prediction equations for various combinations of predictor variables will be made available online for clinical and research use.

## Methods

### Study Population

The present analysis makes use of data from the third wave of follow-up in the Andhra Pradesh Children and Parents Study (APCAPS), in which segmental BIA measurements were obtained. Details of the APCAPS have been published previously^29^. In summary, the study’s baseline data collection took place in 2003-2005 for children whose mothers were involved in the Hyderabad Nutrition Trial from 1987-1990. These children were followed up again in a second wave of data collection in 2009-2010, and the study expanded to also include their siblings and parents during the third wave of data collection in 2010-2012. During this third wave of data collection, a random subset of the study participants were invited for DXA scanning except those who reported for pregnancy. The present analysis is restricted to adults in the APCAPS third wave that consented to DXA scanning, as the second wave of follow-up did not include any BIA measurements.

The APCAPS third wave follow-up study was conducted according to the guidelines laid down in the Declaration of Helsinki and all procedures involving human participants were approved by ethics committees of the Indian Council of Medical Research – National Institute of Nutrition, India (reference number: A2-2009), the Public Health Foundation of India, India (reference number 52/10), and the London School of Hygiene and Tropical Medicine, UK (reference number: 6471). Written informed consent was obtained from all participants (or witnessed thumbprint if illiterate).

### Data Collection

A portable TANITA bioelectrical impedance analyzer (model BC-418 M57NA, TANITA) was used to measure segmental impedance values. To take a measurement, researchers entered the participant’s age, sex, and height into the TANITA machine, then requested participants to stand up straight on the weighing platform in bare feet (with feet in contact with the lower electrodes), while holding onto the hand grips (with hands in contact with the upper electrodes) until a complete impedance reading shows on the display. To reduce measurement variability, readings were taken in the morning following an overnight fast. Height was measured twice to the nearest 1 mm using a portable stadiometer (Leicester height measure), circumferences (hip, waist, calf, head, chest inhaling, chest exhaling, mid arm) were measured twice to the nearest 1 mm using a non-stretch metallic tape measure, skinfolds (tricep, bicep, subscapular, suprailiac, calf) were measured three times to the nearest 0.2 mm using Holtain caliper, and grip strength was measured in kg four times using Lafayette 78010 hand-held dynamometer; the average of each of these measurements were used as a single value in analysis. DXA body composition values were measured using a Hologic Discovery A device with Hologic spine phantom 14855 used as a phantom. The participants were instructed to lay supine with their arms resting by their sides. Standard Hologic software defined the head, trunk, legs, and arms. DXA L1-L4 regions were defined by marking the region from the midpoint of the T12 and L1 vertebrae to the midpoint of the L4 and L5 vertebrae in Hologic software^30^. The total mass, fat mass, and fat percentage were computed within this bounded region for each participant. The L1-L4 scan analyses were performed twice for each participant, and the average of these repeated values was used. This body segment has been used in previous studies as a more valid measure of abdominal fat than trunk fat, since the trunk includes additional body regions such as the chest and pelvis as well as the abdomen^31,32^.

### Data Preparation and Statistical Analysis

Participants were removed from the dataset if they had any missing data or if any of their measurement values were judged to be implausible suggesting machine or data entry error (see Supplementary Material (File 1) for a list of the criteria applied). To account for potential bias/overfitting, the dataset was then split into training and testing groups by randomly selecting rows with 80% of individuals used to fit models and the other 20% used to validate the performance of the models. We explored use of the Least Absolute Selection and Shrinkage Operator (LASSO), random forest, and XGBoost algorithms for developing our prediction equations, as these represent popular parametric (LASSO) and non-parametric, tree-based (random forest, XGBoost) machine learning algorithms. The LASSO model also provides the benefit of automatic feature selection, while random forest and XGBoost allow for more flexible modeling of non-linear relationships between predictors. These algorithms were each used to fit models predicting 6 different DXA-derived outcomes: total body fat (kg), total body lean mass (kg), total body fat percentage (%), trunk fat percentage (%), L1-L4 fat percentage (%), and appendicular lean mass (kg). These six measures were selected as the focus of this paper as they are commonly reported in published research. For completeness, results based on other measures estimated by the DXA machine (trunk fat mass (kg), trunk lean mass (kg), L1-L4 fat mass (kg), L1-L4 lean mass (kg), appendicular fat mass (kg), and appendicular fat mass percentage (%)) are provided in the Supplementary Material (File 4).

The TANITA BC-418 device provides several outputs for the defined body segments of full body, trunk, left arm, right arm, left leg, and right leg. The following measurements as displayed in the TANITA output were used as predictors for each of these segments unless indicated otherwise: segment fat mass, segment fat-free mass, segment muscle mass (all but full body), segment fat percentage, segment impedance. Additionally, body weight, BMI, and total body water were used as predictors. Additional covariates included to potentially improve the predictive performance of the equations were age, standing height, calf circumference, head circumference, exhalation chest circumference, waist circumference, hip circumference, arm circumference, tricep skinfold, bicep skinfold, subscapular skinfold, suprailiac skinfold, calf skinfold, dominant hand grip strength, and non-dominant hand grip strength.

Certain data transformations and interactions were pre-specified for inclusion in the models if deemed to be relevant by the researchers based on review of the literature and potential utility in measuring muscle mass. These interactions and transformations were waist-to-hip ratio, waist-to-height ratio, chest-to-waist ratio, calf-to-height ratio, height-squared-to-impedance ratio for each impedance value, fat mass index (FMI), lean mass index (LMI), body shape index (BSI), corrected arm muscle area (CAMA), logarithm of the sum of skinfolds, and the product of age and dominant hand grip strength. Additionally, we originally included squared terms for each input variable to account for non-linear associations with the outcome and included additional speculative interaction terms to account for complex relationships between predictors. However, the inclusion of these terms did not result in considerable performance improvements, and in some cases worsened model performance due to overfitting. To maintain parsimony while still including potentially meaningful interactions, we used the reduced set of interaction terms described above in the present analysis.

The models were fit separately by sex and performance was compared in the validation data via mean absolute error (MAE), mean average percentage error (MAPE), and root mean squared error (RMSE). RMSE was used to provide comparison to existing studies, while MAE and MAPE were utilized due to their interpretability. The built-in TANITA estimates for each outcome alone were used as a performance baseline. Since TANITA does not define the L1-L4 region, TANITA trunk values were used as the performance baseline for this outcome. Further, we compared the performance of our models to previously derived prediction equations by Kulkarni et al for use in Indian populations for the total body lean mass and appendicular lean mass outcomes^28^ and to Durnin-Womersley equations for total body fat outcomes^26^. The lambda value of the LASSO model, the mtry value of the random forest model, and the max depth, eta, and gamma parameters of the XGBoost model were tuned in the training data using 10-fold cross validation for each outcome.

As a post hoc sensitivity analysis, models were fit using the LASSO algorithm with only the full set of TANITA inputs and relevant transformations, full set of TANITA inputs along with one of the three sets of additional anthropometric variables and their transformations (skinfolds, circumferences, and grip strength), all circumferences alone, and all circumferences and skinfolds and their transformations.

These additional equations and their performance are provided in the Supplementary Material (File 2 and File 3). Age and standing height are still included as predictors in each of these models since it is assumed these values influence the body composition and can be easily obtained by researchers.

All analyses were completed using R software version 4.2.0.

## Results

The details of selecting the study participants are described in a flowchart as Figure 1. The application of inclusion criteria resulted in a sample size of 1615 males and 1422 females (training dataset of 1297 males and 1133 females and validation dataset of 318 males and 289 females). Individuals who did not participate in the DXA study within APCAPS 3^rd^ follow-up tended to be younger on average than individuals who had DXA measurements available (34 years old and 38 years old, respectively), though BMI was similar in the two groups (data not shown). The characteristics of our final study cohort are described in Table 1. Males and females were of similar age (37 years and 38 years old on average, respectively, in both training and test datasets) and with similar BMI (approximately 20-21 kg/m^2^ in all groups). Within the same sex, the average value of each mass outcome (total body fat mass, total body lean mass, appendicular lean mass) did not differ by more than 0.51 kg between training and test datasets and each percentage outcome (total body fat percentage, trunk fat percentage, L1-L4 fat percentage) did not differ by more than 0.75 percentage points between training and test datasets.

**Figure 1:**
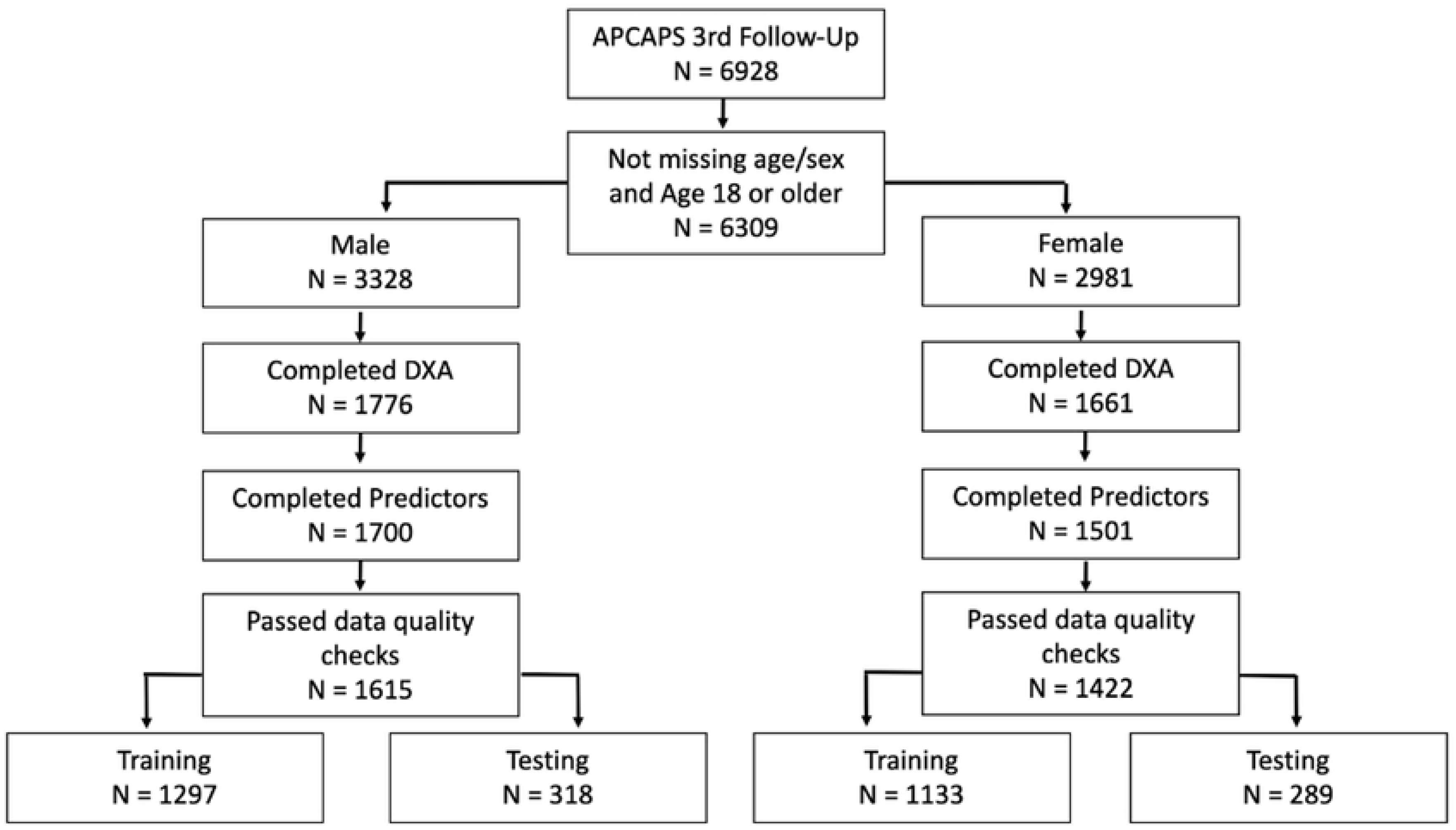
Flow-chart for inclusion in the present analysis.

**Table 1:** Description of the sample.

|  | **Training –**<br>**Females**<br>**(n = 1133)** | **Test –**<br>**Females**<br>**(n = 289)** | **Training –**<br>**Males**<br>**(n = 1297)** | **Test – Males**<br>**(n = 318)** |
| --- | --- | --- | --- | --- |
| **Age (years)** | 38.17 (11.7) | 39.05 (11.3) | 37.15 (15.5) | 36.68 (15.5) |
| **Standing height (cm)** | 151.68 (5.89) | 151.20 (6.05) | 164.26 (6.67) | 164.27 (7.01) |
| **Weight (kg)** | 48.57 (9.20) | 48.39 (9.48) | 55.46 (10.53) | 54.46 (11.42) |
| **BMI (kg/m<sup>2</sup>)** | 21.14 (3.71) | 21.22 (3.85) | 20.55 (3.43) | 20.15 (3.72) |
| **DXA Total body fat mass (kg)** | 15.61 (5.21) | 15.46 (5.25) | 10.68 (4.98) | 10.26 (5.17) |
| **DXA Total body lean mass (kg)** | 32.03 (4.56) | 32.00 (4.95) | 43.52 (6.41) | 43.01 (7.13) |
| **DXA Total body fat percentage (%)** | 30.92 (5.55) | 30.73 (5.72) | 18.25 (5.64) | 17.74 (5.67) |
| **DXA Trunk fat percentage (%)** | 27.02 (6.98) | 26.95 (7.16) | 16.84 (6.56) | 16.09 (6.52) |
| **DXA L1-L4 fat percentage (%)** | 25.80 (7.71) | 25.75 (7.92) | 18.57 (7.69) | 17.83 (7.64) |
| **DXA Appendicular lean mass (kg)** | 13.11 (2.01) | 13.01 (2.25) | 19.18 (2.95) | 19.04 (3.36) |

All values presented as mean (SD)

In general, LASSO models achieved the best performance in the testing data for the 6 outcomes based on each observed performance metric. The performance as measured by MAE of LASSO, random forest, and XGBoost algorithms with the full set of predictors can be found in Table 2. In summary, the LASSO achieved the lowest MAE in the test data for the majority of outcome-sex pairs. The exceptions to this were for prediction of trunk fat percentage in males (MAE of 2.094% by LASSO vs MAE of 2.090% by XGBoost), L1-L4 fat percentage in males (MAE of 2.401% by LASSO vs MAE of 2.369% by random forest), total body fat percentage in females (MAE of 2.165% by LASSO vs MAE of 2.104% by random forest), and trunk fat percentage in females (MAE of 2.721% by LASSO vs MAE of 2.622% by random forest).

**Table 2:** Mean Absolute Error (compared with DXA-based measurement) for different prediction algorithms using all predictors in test data, and additional performance metrics for best model (n = 289 female, 318 male)

| Outcome (Sex) | Random Forest Mean absolute error | XGBoost Mean absolute error | LASSO Mean absolute error | LASSO Root Mean Squared Error | LASSO Mean Absolute Percentage Error |
| --- | --- | --- | --- | --- | --- |
| Total body fat mass (kg) – M | 1.057 | 1.013 | 0.935 | 1.19 | 10.06% |
| Total body fat mass (kg) - F | 1.014 | 1.052 | 0.976 | 1.26 | 6.94% |
| Total body lean mass (kg) - M | 1.311 | 1.378 | 1.177 | 1.52 | 2.76% |
| Total body lean mass (kg) - F | 1.308 | 1.357 | 1.222 | 1.55 | 3.85% |
| Total body fat percentage (%) - M | 1.746 | 1.804 | 1.668 | 2.10 | 9.87% |
| Total body fat percentage (%) - F | 2.104 | 2.125 | 2.165 | 2.73 | 7.44% |
| Trunk fat percentage (%) - M | 2.121 | 2.090 | 2.094 | 2.62 | 13.81% |
| Trunk fat percentage (%) - F | 2.622 | 2.652 | 2.721 | 3.42 | 11.15% |
| L1-L4 fat percentage (%) - M | 2.369 | 2.417 | 2.401 | 2.99 | 14.92% |
| L1-L4 fat percentage (%) - F | 2.971 | 2.970 | 2.929 | 3.75 | 12.41% |
| Appendicular lean mass (kg) - M | 0.900 | 0.853 | 0.782 | 1.04 | 4.18% |
| Appendicular lean mass (kg) - F | 0.833 | 0.860 | 0.790 | 1.00 | 6.17% |

M is male; F is female.

Despite occasional performance gains by the random forest and XGBoost algorithms, we ultimately favor the LASSO algorithm for its comparative interpretability and ease of application to future data. The performance of the LASSO models created for each outcome-sex pair as described by additional performance metrics MAPE and RMSE is also provided in Table 2.

Our novel equations, as well as the other equations previously developed by Kulkarni et al in an Indian population, performed substantially better than the built-in TANITA estimates and Durnin-Womersley equations. For 5 out of the 12 outcome-sex pairs, the mean absolute error of our best equation was less than half that of the TANITA built-in estimate or Durnin-Womersley equation. For example, mean absolute error for total body fat mass was 1.808 kg for males and 2.054 kg for females using TANITA estimation, 2.105 kg for males and 2.995 kg for females using Durnin-Womersley equations, 1.240 kg for males and 1.061 kg for females using our novel equation with just TANITA values, and 0.935 kg for males and 0.976 kg for females using our novel equation with TANITA, skinfolds, circumferences, and grip strength. The mean absolute error for a subset of the LASSO equations (TANITA predictors only, TANITA and skinfold as predictors, and full set of predictors), TANITA built-in estimates, and existing equations as compared to the DXA values are presented in Table 3. In all instances, the best performance in the test data was achieved by our novel equations (built using LASSO algorithm) with all measurements, that is TANITA, circumferences, skinfolds, and grip strength, included. However, similar performance was sometimes achieved through inclusion of only TANITA and skinfolds. The performance metrics for further combinations of measurements are made available in the Supplementary Material (File 3).

**Table 3:** Mean Absolute Error (compared with DXA-based measurement) for the different prediction models in test data (n = 289 female, 318 male)

| Outcome (Sex) | Mean absolute error compared with DXA measurement |  |  |  |  |
| --- | --- | --- | --- | --- | --- |
|  | TANITA Built-in estimate alone | Existing skinfold/ anthropometric equation | All TANITA LASSO | All TANITA + Skinfolks LASSO | All TANITA + Grip strength + circumferences + skinfolks LASSO (I.e., Full LASSO) |
| Total body fat mass (kg) - M | 1.808<sup>a</sup> | 2.105<sup>f</sup> | 1.240 | 1.003 | 0.935 |
| Total body fat mass (kg) - F | 2.054<sup>a</sup> | 2.995<sup>f</sup> | 1.061 | 1.016 | 0.976 |
| Total body lean mass (kg) - M | 2.782<sup>b</sup> | 1.746<sup>g</sup> | 1.401 | 1.230 | 1.177 |
| Total body lean mass (kg) - F | 2.670<sup>b</sup> | 1.402<sup>g</sup> | 1.276 | 1.234 | 1.222 |
| Total body fat percentage (%) - M | 3.242<sup>c</sup> | 3.646<sup>f</sup> | 2.202 | 1.801 | 1.668 |
| Total body fat percentage (%) - F | 4.256<sup>c</sup> | 5.656<sup>f</sup> | 2.298 | 2.193 | 2.165 |
| Trunk fat percentage (%) - M | 3.479<sup>d</sup> | --- | 2.574 | 2.298 | 2.094 |
| Trunk fat percentage (%) - F | 5.837<sup>d</sup> | --- | 2.922 | 2.823 | 2.721 |
| L1-L4 fat percentage (%) - M | 3.883<sup>d</sup> | --- | 2.859 | 2.596 | 2.401 |
| L1-L4 fat percentage (%) - F | 5.494<sup>d</sup> | --- | 3.184 | 3.081 | 2.929 |
| Appendicular lean mass (kg) - M | 1.778<sup>e</sup> | 0.972<sup>g</sup> | 0.926 | 0.826 | 0.782 |
| Appendicular lean mass (kg) - F | 1.548<sup>e</sup> | 0.888<sup>g</sup> | 0.811 | 0.804 | 0.790 |

M is male; F is female. a: TANITA fat mass; b: TANITA fat-free mass; c: TANITA total body fat percentage; d: TANITA trunk fat percentage; e: Sum of TANITA right arm fat-free mass, left arm fat-free mass, right leg fat-free mass, left leg fat-free mass; f: Durnin-Womersley skinfold equations; g: Kulkarni et al Indian-calibrated anthropometric equations.

Briefly, the combination of TANITA and circumferences occasionally led to improved performance from TANITA predictors only, though these performance gains were generally not as large as those achieved by the combination of TANITA and skinfolds. The combination of TANITA and grip strength provided only marginal improvements upon the use of TANITA predictors alone. Bland-Altman plots are provided in Figure 2 to compare predictive performance of our full LASSO equations to that of the TANITA default outputs. Overall, there do not appear to be clear systematic errors in the predictions resulting from our equations, though total fat percentage in women and trunk fat percentage in both men and women may be frequently overestimated by our models among individuals with lower trunk fat percentage. Further, our equations may underestimate appendicular lean mass among women with higher appendicular lean mass. The TANITA device appears to frequently underpredict the fat mass of women with lower body fat mass.

**Figure 2:**
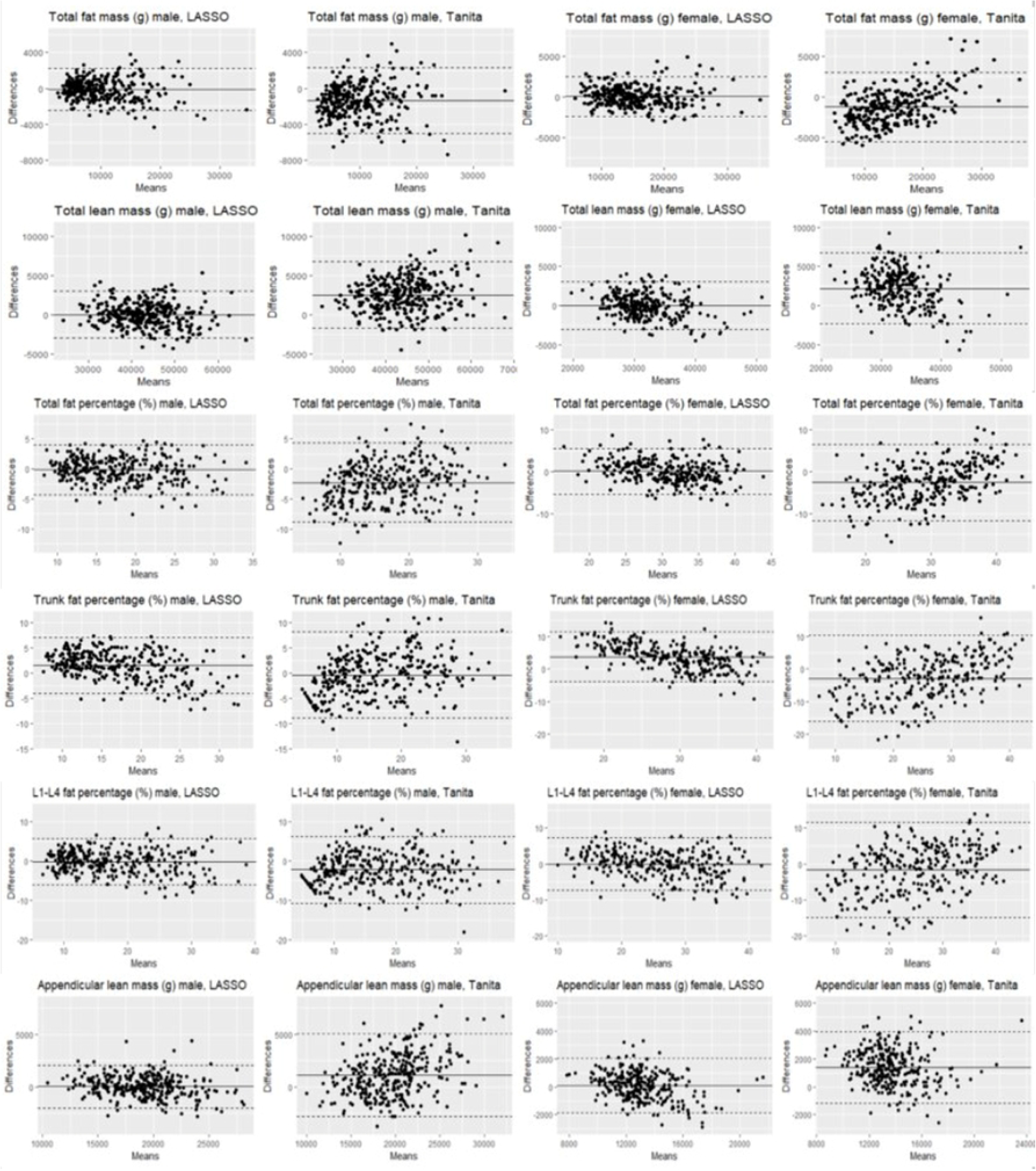
Bland-Altman Plots comparing the LASSO with all variables and TANITA Equivalent Measure alone to DXA values.

The full set of coefficients for LASSO-derived prediction equations are made available online. The coefficients for each outcome-sex pair in the LASSO model with all sets of predictors are provided in the Supplementary Material (File 2). Each row of this table contains an input measure, and each column describes the sex, outcome measure, and intended set of inputs. For example, the column “total fat percentage male Tanita only” provides the set of coefficients for predicting total body fat percentage among males using only the set of TANITA inputs. The predicted value is generated by multiplying each input by its corresponding coefficient from the model of interest and summing together the results (along with the intercept term), as in a standard linear regression model. The coefficient value appears as NA in the table for any predictors that are not used in the corresponding model (e.g. skinfolds in the “Tanita only” model). Estimations can be easily derived from this table using vector multiplication in statistical software. The MAE values for LASSO models with additional sets of predictors are provided in the Supplementary Material (File 3). Performance metrics for additional body composition metrics (e.g. appendicular fat mass) which were not the focus of this manuscript are provided in Supplementary Material (File 4). The 1^st^ and 99^th^ percentiles of each predictor in the training data are provided in Supplementary Material (File 5). Further, all LASSO equations can be used through an interactive application designed by the authors and hosted on GitHub (see data sharing statement).

## Discussion

We have developed and validated the performance of a novel set of equations to predict six different clinically important DXA-derived body composition metrics through low-cost bioelectric impedance analyses calibrated for use with men and women living in and near Hyderabad, India, demonstrating improvement from existing equations developed in other ethnic groups. Accurate measurement of these metrics is important as both fat and lean mass are strong determinants of chronic disease and used clinically in diagnosis, risk stratification, and monitoring of interventions^1–3^. Further, accurate classification of trunk and abdominal fat adiposity may help to illuminate their relationships with cardiometabolic disease among Indian adults, for which direct evidence remains limited^1^. These equations will be particularly useful for large-scale epidemiological studies, especially in settings where expensive DXA scans cannot easily be performed.

Where work by Nigam et al found the mean difference of BIA– and DXA-predicted body fat percentage was 5.4%^10^, our novel model resulted in a mean absolute difference of 1.7% for men and 2.2% for women in testing data. While best performance was achieved by making use of a combination of TANITA, skinfolds, grip strength, and circumferences, our novel equations using only TANITA values as predictors still performed better in the held-out testing data than the TANITA built-in estimates and existing prediction equations for all outcomes. Furthermore, we show that even adding a single measurement to the TANITA values (namely skinfolds, and to a lesser extent circumferences) can improve body composition predictions markedly. This could be useful in resource-constrained contexts where collecting all measurements is not feasible. In addition, we have developed predictive equations for trunk and L1-L4 fat percentage in a cohort of Indian adults, which to our knowledge did not previously exist. The generally improved performance of LASSO as compared to random forest and XGBoost models allows us to provide interpretable prediction equations for wider use. The low-cost nature of the measurements used may be especially beneficial for settings where more expensive and time-consuming methods like DXA and MRI are not feasible (i.e., primary care and community settings).

While we identified several studies validating existing body composition metrics for use in Indian populations, our study represents one of only a few studies developing novel equations to predict DXA-derived values for use with Indian adults. While Dasgupta et al provided similar equations with lower reported RMSE than our study, their study sample consisted only of 117 males aged 18 to 22 years old^25^. As such, the generalizability of these equations may be more limited than the novel equations presented in the current work, developed using a training sample of more than 1000 men and women of a wide age range, the largest population-based DXA dataset including both men and women available in India to our knowledge. The work of Grover et al developed equations to predict DXA-derived fat mass and fat-free mass using BIA, circumferences, and skinfolds separately in a clinical population of 302 adults with cirrhosis but did not present equations using a combination of all factors^33^. The equations by Kulkarni et al utilized skinfolds and waist circumference but did not explore the addition of BIA values into predictive equations^28^. However, the equations by Kulkarni et al resulted in MAE for appendicular lean mass within 0.1 kg of our equations using TANITA only. This is somewhat expected as these equations were calibrated using DXA as reference values and including data from some of the younger individuals in the APCAPS study. Prior development of equations for fat-free mass based on a sample of Black and White individuals by Sun et al reported RMSE of 3.9 kg for males and 2.9 kg for females^8^.

Further, the equations developed by Rush et al for prediction of fat-free mass in an Asian Indian population report an RMSE of 2.13 in the study sample^17^. The RMSE for our LASSO-based equations predicting total lean mass were 1.5 kg for males and 1.6 kg for females, suggesting improved model fit. To our knowledge, the current work represents the introduction of the first set of validated BIA-based equations to predict trunk-specific body composition outcomes for Indian adult men and women.

Though we have demonstrated improved performance from the TANITA output in estimating lean mass in both the full body and appendicular regions, it is unsurprising that the TANITA outputs have a noticeable systematic error. This is because the TANITA uses a two-compartment model to predict fat-free mass, combining both lean mass and skeletal mass into a single measure, whereas the DXA-derived outcome specifically uses a three-compartment model to measure lean mass as distinct from skeletal mass. Additionally, the TANITA provides no direct measure of L1-L4 fat mass as the device cannot detect this body segment. Finally, the TANITA trunk segment includes the mass of the head, while the DXA includes the head as a separate body segment. These observed systematic errors may explain the performance issues of the TANITA observed in our study, though they also emphasize the relevance of the current work. Those interested in measuring lean mass without skeletal mass using TANITA now have a method of doing so despite its use of a two-compartment model.

However, the present work is not without limitations. Of note, not all participants from APCAPS attended the clinic for DXA measurements due to funding limitations or travel distance to the clinic. The participants who did not attend DXA clinics tended to be younger than the participants who did attend the clinics, suggesting there could be systematic differences between those included and those not included due to missing measurements, though no significant difference was found in BMI between the individuals with and without DXA measurements. Regardless, these equations are not recommended for estimating body composition among pregnant women since these women were not included in the sample used for the present development and validation of equations. The validity of these equations in the presence of values outside the range of the training data is also unclear. The training dataset mostly consisted of low-BMI individuals (99^th^ percentile for BMI in both men and women ≈ 31 kg/m^2^). Further, the 99^th^ percentile for age in our data is 67 years in men and 61 years in women. Therefore, predictions may not be suitable for adults with obesity as defined by BMI and those older than the study sample and we recommend interpreting model predictions with caution in such cases. We provide the minimum, 1^st^ percentile, 99^th^ percentile, and maximum of each predictor in the training data in the Supplementary Material (File 5).

Beyond the limitations related to the distribution of variables and selection into the DXA study, limitations to interpretation of results also arise from the use of an internal, rather than external, validation dataset. Our participants are largely from peri-urban areas in southeastern India, and it is unclear how well these equations will perform in other populations in India and in the diaspora, as India is an ethnically diverse country. Additionally, most participants in the APCAPS cohort were between 18-30 or 40-55 years old, meaning that individuals 30-40 years old were underrepresented in the dataset. Because of these limitations, future work is needed to validate these equations in other Indian cohorts and among adults of Indian ethnicity living outside of India. Further, while prior research has shown that DXA is a good substitute for MRI-derived measures of abdominal fat in Indian populations, researchers may still prefer equations developed using MRI for measurement of abdominal fat as the gold standard since DXA may tend to overestimate fat mass in leaner individuals^30^. Additional research may also investigate the performance of these equations against MRI-derived measures of each outcome.

Finally, future validation work is crucial to assess the validity of these equations when deriving BIA estimates from other devices. The present study made use of the TANITA BC-418 device, which although widespread, has since been discontinued by the manufacturer and replaced with the MC-780U. This newer model has been described by the manufacturer as a direct replacement and provides the same 50KHz measurement frequency, suggesting our equations could be used across both devices. Further, the additional measures provided by the new model, such as measurement of impedance using additional frequency values, may present opportunities for development of equations using an expanded set of inputs in the future. Additionally, validation work is needed to assess the performance of these equations when using BIA-based inputs from manufacturers other than TANITA in case of systematic differences between devices. It is the authors’ expectation that the equations will perform well with inputs from any BIA device that can provide segmental estimates, as the underlying approach to impedance estimation is similar, and studies have reported high correspondence in body composition estimates across manufacturers, though one study suggested estimates from Omron devices may differ from other manufacturers^34^. Additionally, it is of note that one study in Finnish adults has demonstrated differences between estimates of fat mass percentage between TANITA and InBody BIA devices, suggesting caution may be needed when using our equations with InBody devices^35^. Our equations are not suitable for use with BIA devices which provide only full-body measurements, as information about the separate body segments are required as input.

## Conclusions

In summary, we have developed and validated several novel prediction equations for a variety of body composition metrics specifically for an Indian population using a large sample size. We encourage the use of our equations based on the LASSO algorithm and make these equations available for use. These equations are expected to deliver improved prediction of various body composition outcomes on average when used with non-pregnant Indian adults. Differences in body composition between different ethnicities due to a combination of factors, including differences between different Asian ethnicities, underscore the importance of developing such equations specifically for adults of Indian ethnicity^36^.

## Declarations

### Ethics approval and consent to participate

The APCAPS third wave follow-up study was conducted according to the guidelines laid down in the Declaration of Helsinki and all procedures involving human participants were approved by ethics committees of the Indian Council of Medical Research – National Institute of Nutrition, India (reference number: A2-2009), the Public Health Foundation of India, India (reference number 52/10), and the London School of Hygiene and Tropical Medicine, UK (reference number: 6471). Written informed consent was obtained from all participants (or witnessed thumbprint if illiterate).

## Consent for publication

Not applicable.

## Availability of data and materials

The APCAPS cohort data analysed in this manuscript are available to bona fide researchers upon request through completion of a collaborator form (https://apcaps.lshtm.ac.uk/apply-to-collaborate/). The equations developed in this manuscript are available through an open-source web application on GitHub (https://github.com/nwbirk).

## Competing interests

The authors declare that they have no competing interest.

## Funding

The Andhra Pradesh Children and Parents’ Study third follow-up was funded by the Wellcome Trust [strategic award 084774]. Authors NB, PACM and SK receive part of their salary from the Medical Research Council (MRC) UK [grant reference MR/V001221/1]. Neither the Wellcome Trust nor the MRC had any role in the design, analysis or writing of this article.

## Authors’ Contributions

NB, PACM and SK conceived the study. NB conducted the study (including analysis of data), wrote the first draft of the manuscript and is responsible for the overall content as guarantor. PACM and SK contributed to study planning. SB, AA, GKW, VG and UR were involved in primary data collection and management. All authors (NB, BK, SB, AA, GKW, VG, UR, HM, SK, PACM) contributed to the writing of subsequent drafts of the manuscript, the interpretation of results, and have given approval for the final version to be published.

## Financial support

The Andhra Pradesh Children and Parents’ Study third follow-up was funded by the Wellcome Trust [strategic award 084774]. Authors NB, PACM and SK receive part of their salary from the Medical Research Council (MRC) UK [grant reference MR/V001221/1]. Neither the Wellcome Trust nor the MRC had any role in the design, analysis or writing of this article.

## Abbreviations

APCAPS: Andhra Pradesh Children and Parents Study
BIA: Bioelectrical impedance analysis
BMI: Body mass index
BSI: Body shape index
CAMA: Corrected arm muscle area
DXA: Dual-energy X-ray absorptiometry
FMI: Fat mass index
LASSO: Least absolute selection and shrinkage operator
LMI: Lean mass index
MAE: Mean absolute error
MAPE: Mean average percentage error
MRI: Magnetic resonance imaging
RMSE: Root mean square error

## Acknowledgments

We are grateful to the participants of the Andhra Pradesh Children and Parents’ Study, the data collection and administrative teams, and the study sponsors. We also gratefully acknowledge the in-kind support of the Indian Council of Medical Research-National Institute of Nutrition.

## Supplementary Material

Supplementary File 1 – List of data quality rules applied for inclusion in the study (Microsoft Word Document)

Supplementary File 2 – Coefficients for each outcome in full model (Microsoft Excel Worksheet)

Supplementary File 3 – Performance (Mean Absolute Error) of the LASSO with alternate sets of predictors (Microsoft Excel Worksheet)

Supplementary File 4 – Information about model performance for other outcomes (trunk fat mass (kg), trunk lean mass (kg), L1-L4 fat mass (kg), L1-L4 lean mass (kg), appendicular fat mass (kg), and appendicular fat mass percentage (%)) (Microsoft Excel Worksheet)

Supplementary File 5 – Minimum, 1st percentile, 99th percentile, and maximum of each predictor in the training data (Microsoft Excel Worksheet)

## Notes

### Competing Interest Statement

The authors have declared no competing interest.

### Funding Statement

Yes

## References

1. Misra A, Khurana L. The Metabolic Syndrome in South Asians: Epidemiology, Determinants, and Prevention. Metab Syndr Relat Disord. 2009;7(6):497–514. doi:10.1089/met.2009.0024

2. Matsuzaki M, Kulkarni B, Kuper H, et al. Association of Hip Bone Mineral Density and Body Composition in a Rural Indian Population: The Andhra Pradesh Children and Parents Study (APCAPS). PloS One. 2017;12(1):e0167114. doi:10.1371/journal.pone.0167114

3. Wolfe RR. The underappreciated role of muscle in health and disease. Am J Clin Nutr. 2006;84(3):475–482. doi:10.1093/ajcn/84.3.475

4. Romero-Corral A, Somers VK, Sierra-Johnson J, et al. Accuracy of body mass index in diagnosing obesity in the adult general population. Int J Obes 2005. 2008;32(6):959–966. doi:10.1038/ijo.2008.11

5. Yajnik CS, Yudkin JS. The Y-Y paradox. Lancet Lond Engl. 2004;363(9403):163. doi:10.1016/S0140-6736(03)15269-5

6. Ward LC. Bioelectrical impedance analysis for body composition assessment: reflections on accuracy, clinical utility, and standardisation. Eur J Clin Nutr. 2019;73(2):194–199. doi:10.1038/s41430-018-0335-3

7. Pietrobelli A, Rubiano F, St-Onge MP, Heymsfield SB. New bioimpedance analysis system: improved phenotyping with whole-body analysis. Eur J Clin Nutr. 2004;58(11):1479–1484. doi:10.1038/sj.ejcn.1601993

8. Sun SS, Chumlea WC, Heymsfield SB, et al. Development of bioelectrical impedance analysis prediction equations for body composition with the use of a multicomponent model for use in epidemiologic surveys. Am J Clin Nutr. 2003;77(2):331–340. doi:10.1093/ajcn/77.2.331

9. Haroun D, Taylor SJC, Viner RM, et al. Validation of bioelectrical impedance analysis in adolescents across different ethnic groups. Obes Silver Spring Md. 2010;18(6):1252–1259. doi:10.1038/oby.2009.344

10. Nigam P, Misra A, Colles SL. Comparison of DEXA-derived body fat measurement to two race-specific bioelectrical impedance equations in healthy Indians. Diabetes Metab Syndr. 2013;7(2):72–77. doi:10.1016/j.dsx.2013.02.031

11. Kuriyan R, Thomas T, Ashok S, Jayakumar J, Kurpad AV. A 4-compartment model based validation of air displacement plethysmography, dual energy X-ray absorptiometry, skinfold technique & bio-electrical impedance for measuring body fat in Indian adults. Indian J Med Res. 2014;139(5):700–707.

12. Bhat DS, Yajnik CS, Sayyad MG, et al. Body fat measurement in Indian men: comparison of three methods based on a two-compartment model. Int J Obes 2005. 2005;29(7):842–848. doi:10.1038/sj.ijo.0802953

13. Kehoe SH, Krishnaveni GV, Lubree HG, et al. Prediction of body-fat percentage from skinfold and bio-impedance measurements in Indian school children. Eur J Clin Nutr. 2011;65(12):1263–1270. doi:10.1038/ejcn.2011.119

14. Iyengar A, Kuriyan R, Kurpad AV, Vasudevan A. Body Fat in Children with Chronic Kidney Disease – A Comparative Study of Bio-impedance Analysis with Dual Energy X-ray Absorptiometry. Indian J Nephrol. 2021;31(1):39–42. doi:10.4103/ijn.IJN_368_19

15. Chiplonkar S, Kajale N, Ekbote V, et al. Validation of Bioelectric Impedance Analysis against Dual-energy X-Ray Absorptiometry for Assessment of Body Composition in Indian Children Aged 5 to 18 Years. Indian Pediatr. 2017;54(11):919–924. doi:10.1007/s13312-017-1182-3

16. Deurenberg P, Deurenberg-Yap M, Schouten F. Validity of total and segmental impedance measurements for prediction of body composition across ethnic population groups. Eur J Clin Nutr. 2002;56(3):214–220. doi:10.1038/sj.ejcn.1601303

17. Rush EC, Chandu V, Plank LD. Prediction of fat-free mass by bioimpedance analysis in migrant Asian Indian men and women: a cross validation study. Int J Obes 2005. 2006;30(7):1125–1131. doi:10.1038/sj.ijo.0803230

18. Rush EC, Freitas I, Plank LD. Body size, body composition and fat distribution: comparative analysis of European, Maori, Pacific Island and Asian Indian adults. Br J Nutr. 2009;102(4):632–641. doi:10.1017/S0007114508207221

19. Marwaha RK, Tandon N, Garg MK, Narang A, Mehan N, Bhadra K. Normative data of body fat mass and its distribution as assessed by DXA in Indian adult population. J Clin Densitom Off J Int Soc Clin Densitom. 2014;17(1):136–142. doi:10.1016/j.jocd.2013.01.002

20. Hunma S, Ramuth H, Miles-Chan JL, et al. Body composition-derived BMI cut-offs for overweight and obesity in Indians and Creoles of Mauritius: comparison with Caucasians. Int J Obes 2005. 2016;40(12):1906–1914. doi:10.1038/ijo.2016.176

21. Shah AD, Kandula NR, Lin F, et al. Less favorable body composition and adipokines in South Asians compared with other US ethnic groups: results from the MASALA and MESA studies. Int J Obes. 2016;40(4):639–645. doi:10.1038/ijo.2015.219

22. Kapoor N. Thin Fat Obesity: The Tropical Phenotype of Obesity. In: Feingold KR, Anawalt B, Boyce A, et al., eds. Endotext. MDText.com, Inc.; 2000. Accessed January 12, 2023. http://www.ncbi.nlm.nih.gov/books/NBK568563/

23. Kapoor N, Furler J, Paul TV, Thomas N, Oldenburg B. Normal Weight Obesity: An Underrecognized Problem in Individuals of South Asian Descent. Clin Ther. 2019;41(8):1638–1642. doi:10.1016/j.clinthera.2019.05.016

24. Bhopal RS. Epidemic of Cardiovascular Disease and Diabetes: Explaining the Phenomenon in South Asians Worldwide. First edition. Oxford University Press; 2019.

25. Dasgupta R, Anoop S, Samuel P, et al. Bioimpedance analysis with a novel predictive equation – A reliable technique to estimate fat free mass in birth weight based cohorts of Asian Indian males. Diabetes Metab Syndr. 2019;13(1):738–742. doi:10.1016/j.dsx.2018.11.070

26. Durnin JV, Womersley J. Body fat assessed from total body density and its estimation from skinfold thickness: measurements on 481 men and women aged from 16 to 72 years. Br J Nutr. 1974;32(1):77–97. doi:10.1079/bjn19740060

27. Davidson LE, Wang J, Thornton JC, et al. Predicting Fat Percent by Skinfolds in Racial Groups: Durnin and Womersley Revisited. Med Sci Sports Exerc. 2011;43(3):542–549. doi:10.1249/MSS.0b013e3181ef3f07

28. Kulkarni B, Kuper H, Taylor A, et al. Development and validation of anthropometric prediction equations for estimation of lean body mass and appendicular lean soft tissue in Indian men and women. J Appl Physiol Bethesda Md 1985. 2013;115(8):1156–1162. doi:10.1152/japplphysiol.00777.2013

29. Kinra S, Radha Krishna KV, Kuper H, et al. Cohort profile: Andhra Pradesh Children and Parents Study (APCAPS). Int J Epidemiol. 2014;43(5):1417–1424. doi:10.1093/ije/dyt128

30. Taylor AE, Kuper H, Varma RD, et al. Validation of dual energy X-ray absorptiometry measures of abdominal fat by comparison with magnetic resonance imaging in an Indian population. PloS One. 2012;7(12):e51042. doi:10.1371/journal.pone.0051042

31. Glickman SG, Marn CS, Supiano MA, Dengel DR. Validity and reliability of dual-energy X-ray absorptiometry for the assessment of abdominal adiposity. J Appl Physiol Bethesda Md 1985. 2004;97(2):509–514. doi:10.1152/japplphysiol.01234.2003

32. Paradisi G, Smith L, Burtner C, et al. Dual energy X-ray absorptiometry assessment of fat mass distribution and its association with the insulin resistance syndrome. Diabetes Care. 1999;22(8):1310–1317. doi:10.2337/diacare.22.8.1310

33. Grover I, Singh N, Gunjan D, Pandey RM, Chandra Sati H, Saraya A. Comparison of Anthropometry, Bioelectrical Impedance, and Dual-energy X-ray Absorptiometry for Body Composition in Cirrhosis. J Clin Exp Hepatol. 2022;12(2):467–474. doi:10.1016/j.jceh.2021.05.012

34. Alivia Blakely. Validity of Various Bioelectrical Impedance Analysis Devices vs the Bod Pod for Body Composition. Masters. Cleveland State University; 2019. https://engagedscholarship.csuohio.edu/etdarchive/1161/

35. Volgyi E, Tylavsky FA, Lyytikainen A, Suominen H, Alen M, Cheng S. Assessing Body Composition With DXA and Bioimpedance: Effects of Obesity, Physical Activity, and Age. Obesity. 2008;16(3):700–705. doi:10.1038/oby.2007.94

36. Deurenberg P, Deurenberg-Yap M, Guricci S. Asians are different from Caucasians and from each other in their body mass index/body fat per cent relationship. Obes Rev Off J Int Assoc Study Obes. 2002;3(3):141–146. doi:10.1046/j.1467-789x.2002.00065.x

